# Serial SARS-CoV-2 antibody titers in vaccinated dialysis patients: prevalence of unrecognized infection and duration of seroresponse

**DOI:** 10.1101/2023.03.16.23287322

**Authors:** Caroline M. Hsu, Daniel E. Weiner, Harold J. Manley, Dana Miskulin, Vladimir Ladik, Jill Frament, Christos Argyropoulos, Kenneth Abreo, Andrew Chin, Reginald Gladish, Loay Salman, Doug Johnson, Eduardo K. Lacson

**Affiliations:** Tufts Medical Center, Boston, MA; Dialysis Clinic Inc., Nashville, TN; University of New Mexico, Albuquerque, NM; Louisiana State University Health Sciences Center, Shreveport, LA; University of California, Davis, Sacramento, CA; Nephrology of North Alabama, Decatur, AL; Albany Medical College, Albany, NY

**Author notes:** **Address for Correspondence:** Caroline M. Hsu, 800 Washington St, Box #391, Boston, MA 02111.

## Abstract

**Rationale & Objective:** SARS-CoV-2 infections are likely underdiagnosed, but the degree of underdiagnosis among maintenance dialysis patients is unknown. Durability of the immune response after third vaccine doses in this population also remains uncertain. This study tracked antibody levels to 1) assess the rate of undiagnosed infections and 2) characterize seroresponse durability after third doses.

**Study Design:** Retrospective observational study

**Setting & Participants:** SARS-CoV-2 vaccinated patients receiving maintenance dialysis through a national dialysis provider. Immunoglobulin G spike antibodies (anti-spike IgG) titers were assessed monthly following vaccination.

**Exposure(s):** Two and three doses of SARS-CoV-2 vaccine

**Outcome(s):** Undiagnosed and diagnosed SARS-CoV-2 infections; anti-spike IgG titers over time

**Analytical Approach:** “Undiagnosed” SARS-CoV-2 infections were identified as an increase in anti-spike IgG titer of ≥ 100 BAU/mL, not associated with receipt of vaccine or “diagnosed” SARS-CoV-2 infection (by PCR or antigen test). In descriptive analyses, anti-spike IgG titers were followed over time.

**Results:** Among 2660 patients without prior COVID-19 who received an initial two-dose vaccine series, 371 (76%) SARS-CoV-2 infections were diagnosed and 115 (24%) were undiagnosed. Among 1717 patients without prior COVID-19 who received a third vaccine dose, 155 (80%) SARS-CoV-2 infections were diagnosed and 39 (20%) were undiagnosed. In both cohorts, anti-spike IgG levels declined over time. Of the initial two-dose cohort, 66% had a titer ≥ 500 BAU/mL in the first month, with 23% maintaining a titer ≥ 500 BAU/mL at six months. Of the third dose cohort, 95% had a titer ≥ 500 BAU/mL in the first month after the third dose, with 76% maintaining a titer ≥ 500 BAU/mL at six months.

**Limitations:** Assays used had upper limits.

**Conclusions:** Among maintenance dialysis patients, 20-24% of SARS-CoV-2 infections were undiagnosed. Given this population’s vulnerability to COVID-19, ongoing infection control measures are needed. A three-dose primary mRNA vaccine series optimizes seroresponse rate and durability.

## Introduction

Patients receiving maintenance dialysis are particularly vulnerable to poor outcomes from COVID-19, due to immunocompromised state, limited ability to physically distance, and high comorbid burden.^1^ Studies have shown that the majority of dialysis patients mount a robust immune response to mRNA vaccines,^2,3^ but this immune response is weaker and wanes more quickly than in the healthy adult population.^4–6^ Additional doses of vaccine augment the immune response, including among those who failed to mount an immune response initially.^7,8^ Higher antibody response has been associated with a reduced incidence of breakthrough infection and reduced morbidity of those infected.^5,9^

SARS-CoV-2 infections likely are underdiagnosed, potentially reflecting milder symptoms related to immunity induced by vaccines or prior infections and, potentially, less virulence of evolving strains.^10^ The degree of underdiagnosis has not been investigated among maintenance dialysis patients, a particularly unique population in both their risk for exposure and their opportunities for diagnosis, given the frequency of contact with the healthcare system. In addition, the durability of the immune response after additional (third) doses and booster vaccine doses among maintenance dialysis patients remains uncertain. The initial augmentation of the immune response by additional vaccine doses is well-established,^7,8^ but longer term studies have been limited by small sample size and infrequent monitoring.^11,12^ In this study, we used serially collected antibody data to 1) identify previously unrecognized SARS-CoV-2 infections based on an increase in titer, and 2) characterize the trend of antibody titer levels over six months after additional vaccine doses.

## Methods

### Antibody assessment

Dialysis Clinic, Inc. (DCI) is a national not-for-profit dialysis provider that cares for approximately 15,000 patients at 260 outpatient dialysis clinics across 29 states. As previously described,^4^ since January 2021 physicians at DCI facilities have had available an antibody monitoring protocol for patients, activated by physician order upon documentation of receipt of a SARS-CoV-2 vaccine, regardless of the vaccine type or place of administration. Like the existing hepatitis B vaccine protocol, the SARS-CoV-2 vaccine protocol documents seroresponse to vaccination by measuring antibody titers as part of the monthly blood draws. The assay measures immunoglobulin G spike antibodies (anti-spike IgG) against the receptor-binding domain of the S1 subunit of SARS-CoV-2 spike antigen. The ADVIA Centaur® XP/XPT COV2G assay was used from January 1, 2021 to September 30, 2021, and the manufacturer-updated ADVIA Centaur® sCOVG assay was used from October 1, 2021 onwards. Both are chemiluminescent semi-quantitative assays that report an Index Value established with calibrators. These Index aalues were then converted to binding antibody units per mL (BAU/mL)^13^, with the COV2G assay providing values between 0 and ≥ 837 BAU/mL and the sCOVG assay providing values between 0 and ≥ 1781 BAU/mL. The manufacturer recommends a threshold equivalent to 45 BAU/mL as representing seropositivity. The handling of values at the upper limit of the assays is described throughout the specific analyses. Data were censored at transplant, kidney recovery, or April 30, 2022, whichever occurred first. If a patient had more than one titer assessed in a calendar month, only the first measurement was retained. Handling of missing values is described in the **Supplemental Methods**.

This study was reviewed and approved by the WCG IRB Work Order 1-1456342-1. Statistical analyses were performed using R v4.0.2.

### Population

Demographic and clinical data, vaccination dates, and anti-spike IgG titer results were obtained from the DCI electronic health record. This study includes adult patients (age ≥ 18 years). Patients within the “initial series” cohort could be included in the “third dose” subcohort upon receipt of a third dose of SARS-CoV-2 vaccine. The “initial series cohort” includes all patients who were fully vaccinated (≥ 14 days after two mRNA-1273/Moderna vaccine doses or two BNT162b2/Pfizer vaccine doses or one Ad26.COV2.S/Janssen vaccine dose) and had at least one anti-spike IgG titer assessment after full vaccination by this initial vaccine series and prior to any subsequent clinical COVID-19 diagnosis or receipt of a third dose. The “third dose” subcohort is the subset of the “initial series” cohort who received an additional vaccine dose and had at least one anti-spike IgG titer assessment after receipt of the additional vaccine dose and prior to any subsequent clinical COVID-19 diagnosis or receipt of a fourth dose. An additional vaccine dose is defined as a third dose for those who received a two-dose Moderna or Pfizer series initially or as a second dose for those who received a Janssen dose initially; because the vast majority of included patients received either the Moderna or Pfizer vaccine series initially, we use the notation “third dose” for clarity. For patients who received a fourth dose of vaccine, the handling of these doses is described in the Analytic Plan below. Analyses were conducted in parallel on each of these cohorts.

### Identification of SARS-CoV-2 infections based on an increase in titer

To minimize the effects of assay variability, a 3-month rolling average of the titers was used for these analyses. We defined “undiagnosed SARS-CoV-2 infection” as an increase of 100 BAU/mL or more from one rolling average to the next, not including any increase occurring after or up to seven days before a “diagnosed SARS-CoV-2 infection,” defined as any positive PCR or antigen test. This definition of undiagnosed SARS-CoV-2 infection also excludes any increase occurring within 60 days after completing an initial two-dose mRNA vaccine series, within 90 days after an initial Janssen dose, or within 60 days after receiving an additional (third) vaccine dose. The selection of 100 BAU/mL as a threshold is described in the **Supplemental Methods**, with a 200 BAU/mL threshold used in sensitivity analyses.

Of note, all DCI patients are screened for COVID-19 symptoms and recent exposure upon arrival to the dialysis facility for each hemodialysis treatment or for each peritoneal dialysis encounter, followed by SARS-CoV-2 testing if they screen positive. Positive SARS-CoV-2 tests were captured regardless of where the patient was tested (e.g., in the dialysis clinic, at a testing center, at a hospital, at home), and great care was taken to capture testing results at all sites as this information determines patients’ need for isolation during dialysis treatment. Patients were assessed for the first of diagnosed SARS-CoV-2 infection or undiagnosed SARS-CoV-2 infection.

### Analytic Plan

Study entry is defined by vaccination against SARS-CoV-2. Analyses were stratified by whether a patient had COVID-19 prior to receipt of vaccine. Prior COVID-19 status was defined by a positive SARS-CoV-2 PCR test at any time before the date of full vaccination or anti-spike IgG titer ≥ 45 BAU/mL before or within 10 days after the first vaccine dose (representing likely undiagnosed COVID-19 at the time of initial vaccine receipt, as suggested by prior studies^14,15^). Analyses were further grouped by vaccine type. Patients were identified as developing diagnosed or undiagnosed SARS-CoV-2 infections during the post-vaccination period as described above.

#### Initial series cohort

Analyses of this cohort followed antibody titer levels after completion of the initial series. As a result, for most patients these levels spanned the change in assay described above. To account for this change, analyses placed a limit at ≥ 837 BAU/mL, the upper limit of the COV2G assay, which was used initially. To assess trends over time, descriptive analyses compared titers by vaccine type. Anti-spike IgG titers were grouped by the month of assessment relative to the date of full vaccination (month 1, 2, etc…). These analyses censored titers at the time of SARS-CoV-2 infection (whether diagnosed or undiagnosed), or at receipt of a third vaccine dose.

#### Third dose subcohort

Analyses were stratified by prior COVID-19 status and were further grouped by the type of third (additional) vaccine dose received. Analyses of this cohort followed their antibody titer levels from receipt of the third vaccine dose. Due to the timing of the rollout of third doses, nearly all titers after the third dose were collected after the assay change described above. Therefore analyses are shown using the sCOVG assay’s full range, from 0 to ≥ 1781 BAU/mL. The titers at the upper limit of the COV2G assay were adjudicated with the upper limit of the sCOVG assay, ≥ 1781 BAU/mL; the reasoning for this is in the **Supplemental Methods**.

To assess antibody titer trends over time, descriptive analyses compared titers by type of third vaccine dose. Anti-spike IgG titers were grouped by the month of assessment relative to 14 days after receipt of a third dose (month 1, 2, etc). These analyses censored titers at SARS-CoV-2 infection (whether diagnosed or undiagnosed), or at receipt of a second additional (fourth) dose.

## Results

Among DCI patients receiving maintenance dialysis, 4183 adults received a full SARS-CoV-2 vaccine series and had at least one anti-spike IgG titer assessment. Of these, 3171 (76%) had at least one anti-spike IgG titer assessment after receipt of an initial vaccine series and prior to any subsequent COVID-19 diagnosis or receipt of a third dose. Of the 4183 patients who received a full, two-dose SARS-CoV-2 vaccine series (or a single dose of the Janssen vaccine), 2800 (67%) received a third dose. Of these, 2005 (72%) had at least one anti-spike IgG titer assessment after their third dose and prior to any subsequent COVID-19 diagnosis or receipt of a fourth vaccine dose (**Figure 1)**.

**Figure 1.**
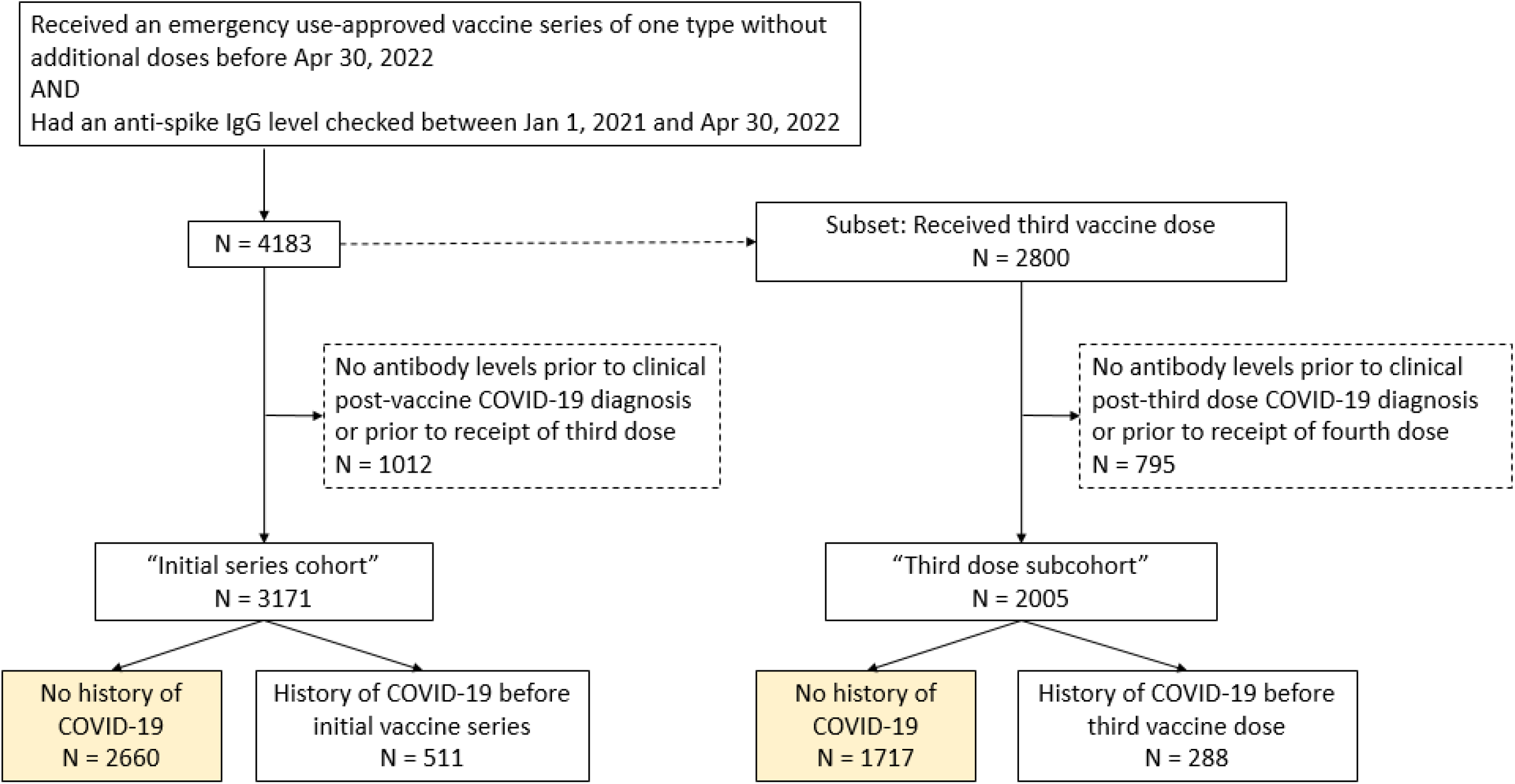
Consort diagram.

### Initial vaccine series cohort

Among the 3171 patients who completed an initial vaccine series, the mean age was 64 ± 14 (SD) years, 1828 (58%) were male, and 153 (5%) resided in a long-term care facility. (**Table 1)** Among the 511 (16%) patients who had COVID-19 prior to full immunization by the initial vaccine series, titers largely remained at the upper limit of the assay (**Supplemental Figure 1**). For this reason, identification of infection by increase in antibody titer could not be conducted among those with prior COVID-19.

**Table 1.** Baseline characteristics.

|  | Initial series cohort |  | Third dose subcohort |  |
| --- | --- | --- | --- | --- |
|  | No prior COVID-19<br>N = 2660 | Prior COVID-19<br>N = 511 | No prior COVID-19<br>N = 1717 | Prior COVID-19<br>N = 288 |
| Age | 64.2 ± 14.1 | 61.1 ± 13.6 | 64.8 ± 13.2 | 61.9 ± 13.6 |
| Male sex | 1535 (57.7) | 293 (57.3) | 1023 (59.6) | 166 (57.6) |
| Race |  |  |  |  |
| Native American | 173 (6.5) | 58 (11.4) | 121 (7.0) | 30 (10.4) |
| Asian/Pacific Islander | 124 (4.7) | 18 (3.5) | 80 (4.7) | 19 (6.6) |
| Black | 622 (23.4) | 128 (25.0) | 364 (21.2) | 76 (26.4) |
| Unknown/Other | 332 (12.5) | 50 (9.8) | 272 (15.8) | 25 (8.7) |
| White | 1409 (53.0) | 257 (50.3) | 880 (51.3) | 138 (47.9) |
| Hispanic ethnicity | 275 (10.3) | 112 (21.9) | 225 (13.1) | 67 (23.3) |
| Vintage, months | 30.7 [10.2, 63.5] | 37.7 [17.8, 67.8] | 28.5 [7.2, 60.9] | 40.2 [18.5, 74.6] |
| Body mass index, kg/m<sup>2</sup> | 29.0 ± 7.3 | 29.0 ± 7.2 | 29.2 ± 7.4 | 29.2 ± 7.2 |
| Diabetes | 1506 (56.6) | 331 (64.8) | 976 (56.8) | 199 (69.1) |
| Long-term care facility | 92 (3.5) | 61 (11.9) | 44 (2.6) | 35 (12.2) |
| Modality |  |  |  |  |
| Home hemodialysis | 56 (2.1) | 2 (0.4) | 33 (1.9) | 3 (1.0) |
| In-center hemodialysis | 2268 (85.3) | 466 (91.2) | 1461 (85.1) | 273 (94.8) |
| Peritoneal dialysis | 336 (12.6) | 43 (8.4) | 223 (13.0) | 12 (4.2) |
| Albumin, g/dL | 3.9 ± 0.4 | 3.8 ± 0.4 | 3.9 ± 0.4 | 3.9 ± 0.4 |
| HBsAb ≥10 mIU/mL <sup>a</sup> | 755 (28.4) | 183 (35.8) | 452 (26.3) | 99 (34.4) |
| History of transplantation | 103 (3.9) | 17 (3.3) | 79 (4.6) | 9 (3.1) |
| Immunodeficiency | 470 (17.7) | 79 (15.5) | 310 (18.1) | 38 (13.2) |
| Immunomodulating medication <sup>b</sup> | 343 (12.9) | 63 (12.3) | 208 (12.1) | 33 (11.5) |
| Congestive heart failure | 472 (17.7) | 83 (16.2) | 269 (15.7) | 40 (13.9) |
| Peripheral vascular disease | 265 (10.0) | 56 (11.0) | 136 (7.9) | 22 (7.6) |
| Cerebrovascular disease | 192 (7.2) | 42 (8.2) | 109 (6.3) | 26 (9.0) |
| Chronic obstructive pulmonary disease | 353 (13.3) | 60 (11.7) | 189 (11.0) | 28 (9.7) |
| History of cancer | 226 (8.5) | 44 (8.6) | 140 (8.2) | 23 (8.0) |
| Duration of follow-up, days | 210 [154, 271] | 210 [146, 289] | 134 [84, 181] | 141 [87, 180] |
| Time between initial vaccine series and third dose, days | --- | --- | 174 [140, 205] | 168 [125, 204] |

Vintage and duration of follow-up are reported as median [IQR]. All other data are reported as mean ± standard deviation or %.
Data on baseline patient characteristics were complete.
<sup>a</sup> HBsAb ≥10 mIU/mL signifies hepatitis B seroimmunity
<sup>b</sup> Immunomodulating medications include anti-inflammatory medications, anti-neoplastic agents, corticosteroids, and certain anti-infective medications

Among the 2660 patients without a prior history of COVID-19, 486 (18%) had a SARS-CoV-2 infection; 115 (24%) were clinically undiagnosed and reflected only with antibody titer levels while 371 (76%) were diagnosed clinically. A greater number of undiagnosed infections occurred in the summer of 2021 and the winter of 2021-2022, overlapping with the Delta and Omicron waves in the United States, respectively (**Figure 2**).^16^ The association of vaccine type with diagnosed and undiagnosed SARS-CoV-2 infections is shown in **Table 2**. Janssen vaccine recipients had the highest rate of total SARS-CoV-2 infections. Moderna and Pfizer vaccine recipients had similar rates of total SARS-CoV-2 infections, but Pfizer vaccine recipients had a higher rate of diagnosed infections. In sensitivity analyses using a threshold of an increase of 200 BAU/mL, there were 85 undiagnosed infections identified, a reduction of 30 compared to using the 100 BAU/mL threshold (**Supplemental Table 1**).

**Figure 2.**
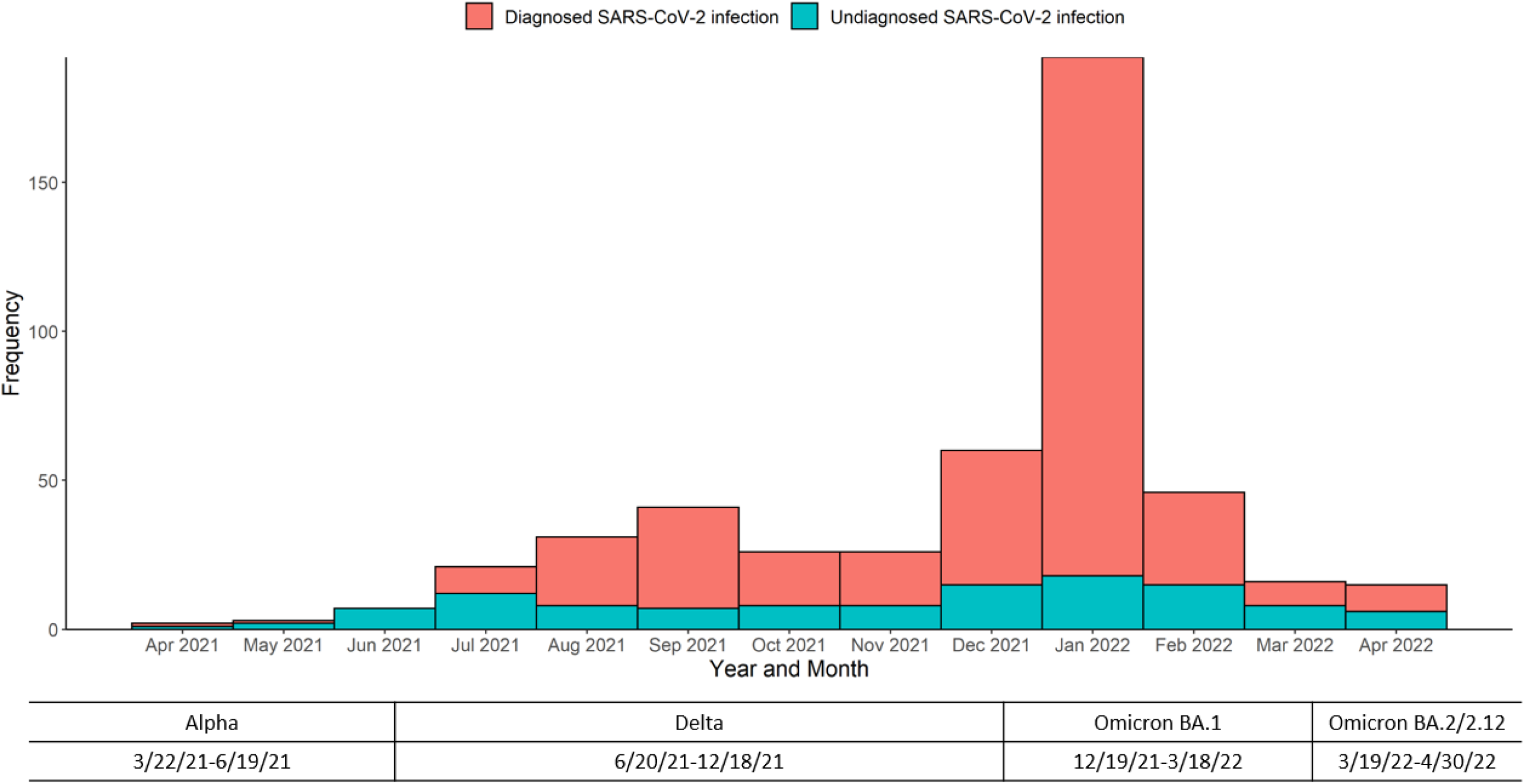
Timing of titer-identified SARS-CoV-2 infections, initial series cohort.

**Table 2.** Association of vaccine type with diagnosed and undiagnosed SARS-CoV-2 infections within the initial vaccine series cohort, among those without history of COVID-19.

|  | **Total** | Ad26.COV2.S/<br>Janssen | mRNA-1273/<br>Moderna | BNT162b2/<br>Pfizer |
| --- | --- | --- | --- | --- |
| Diagnosed SARS-CoV-2 infection | **371 (14%)** | 74 (19%) | 169 (12%) | 128 (16%) |
| Undiagnosed SARS-CoV-2 infection | **115 (4%)** | 34 (9%) | 58 (4%) | 23 (3%) |
| No SARS-CoV-2 infection | **2174 (82%)** | 286 (73%) | 1224 (84%) | 664 (81%) |
| Total | **2660** | 394 | 1451 | 815 |

p < 0.01 by Chi-square test

Antibody titers were trended over time. Among mRNA vaccine recipients, titers waned over time among those without a prior history of COVID-19 (**Figure 3A**). Among Moderna vaccine recipients, median [IQR] titers were ≥ 837 [≥ 837, ≥ 837] in month 1, 276 [117, ≥ 837] in month 6, and 209 [66, ≥ 837] in month 12. Among Pfizer vaccine recipients, median [IQR] titers were ≥ 837 [386, ≥ 837] in month 1, 120 [44, 256] in month 6, and 0 [0, 271] in month 12. Only 33% of Moderna vaccine recipients and 16% of Pfizer vaccine recipients still had a titer of at least 500 BAU/mL at six months after the initial series in the absence of infection.

**Figure 3.**
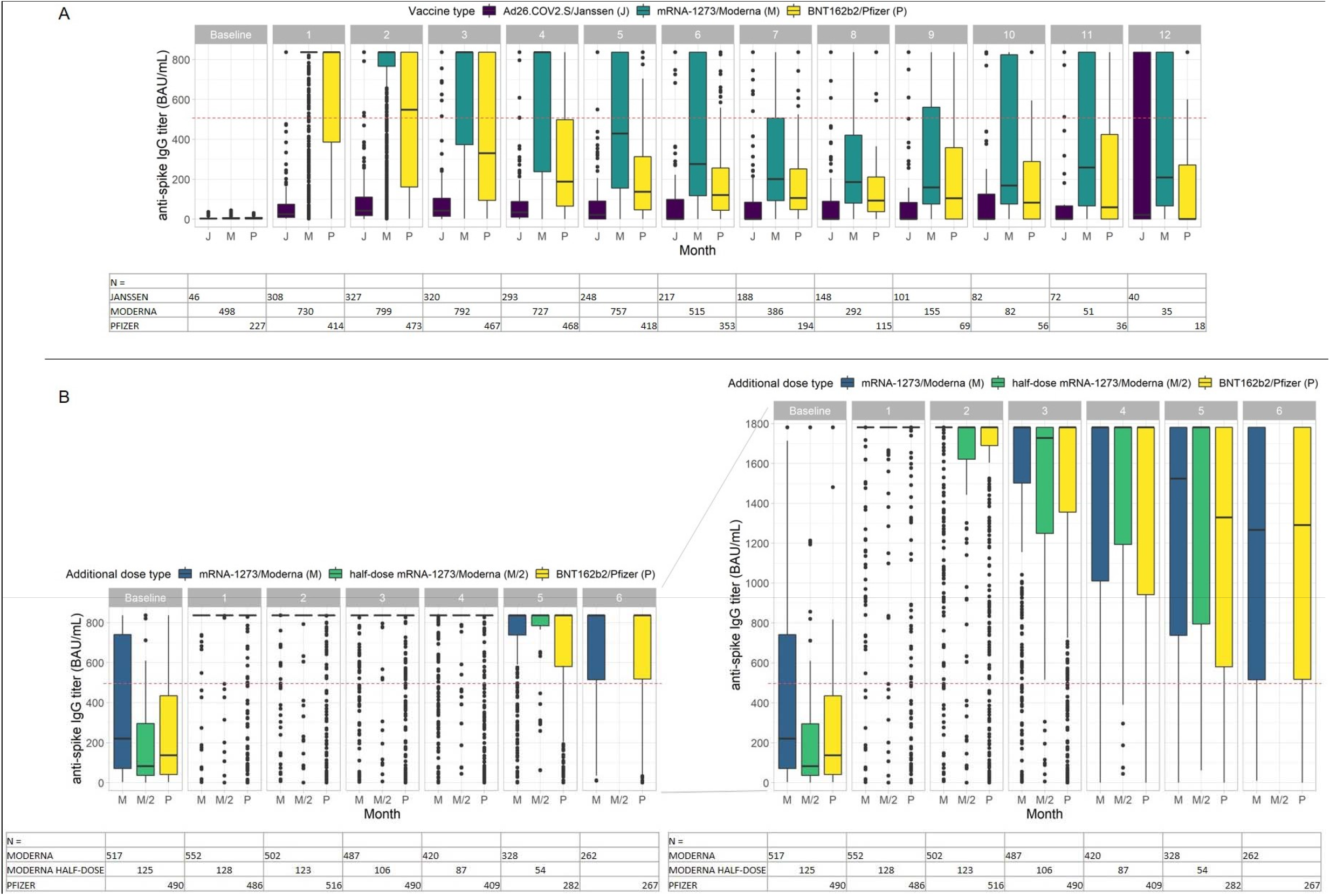
Anti-spike IgG titers vs months (A) after date of full immunization by initial vaccine series, and (B) after date of full immunization by third vaccine dose, among patients without prior COVID-19. The boxplots shown here are bounded by the upper and lower limits of the assay used. For example, in (A), among the mRNA-1273/Moderna recipients, the median [IQR] titer was ≥ 837 [≥ 837, ≥ 837] in month 1 and ≥ 837 [374, ≥ 837] in month 3. In (B), among the mRNA-1273/Moderna recipients, the median [IQR] titer was ≥ 1781 [≥ 1781, ≥ 1781] in month 1 and ≥ 1781 [1502, ≥ 1781] in month 3; this is shown on the left capped at ≥ 837 to enable comparison with (A), and it is shown on the right with the updated assay’s full range. The dots represent the outliers, defined as greater than 1.5*IQR above the third quartile or less than 1.5*IQR below the first quartile. The tables of N show the number of titers for each month, by vaccine type. There were only two recipients of the Moderna half-dose in month 6 in (B), so their data are not displayed.

### Third (additional) dose cohort

Among 2005 patients who received a third (additional) vaccine dose, the average age was 64 ± 13 (SD) years, 1189 (59%) were male, and 79 (4%) lived in a long-term care facility. (**Table 1**) The breakdown of vaccine type received, by initial vaccine series and by third dose, is shown in **Supplemental Table 2**. Among the 288 (14%) patients who had COVID-19 prior to the third dose, 5 recipients of the Janssen vaccine as a third dose and 17 recipients of the half-dose Moderna vaccine as a third vaccine dose were excluded from analysis due to small sample size. Among the remaining 266 patients, titers largely remained at the upper limit of the assay (**Supplemental Figure 2**); accordingly, identification of infection by increase in antibody titer could not be conducted among those with prior COVID-19.

Among the 1717 patients without a prior history of COVID-19, 194 (11%) had a SARS-CoV-2 infection; 39 (20%) were undiagnosed and based on antibody titer levels and 155 (80%) were diagnosed. Many of the otherwise unexplained increases in antibody titer levels occurred from late 2021 into spring of 2022, coinciding with the Omicron wave in the United States (**Figure 4**).^16^ The association of vaccine type (type of initial series and type of third dose) with diagnosed and undiagnosed infections is shown in **Table 3**, and there was little difference by vaccine type. Although recipients of Janssen vaccine as a third dose had the highest rate of total SARS-CoV-2 infections, there were only 19 Janssen vaccine recipients in total, compared to 1698 mRNA vaccine recipients in total. In sensitivity analyses using a threshold of an increase of 200 BAU/mL, there were 23 undiagnosed infections identified, a reduction of 16 compared to using the 100 BAU/mL threshold (**Supplemental Table 1**).

**Figure 4.**
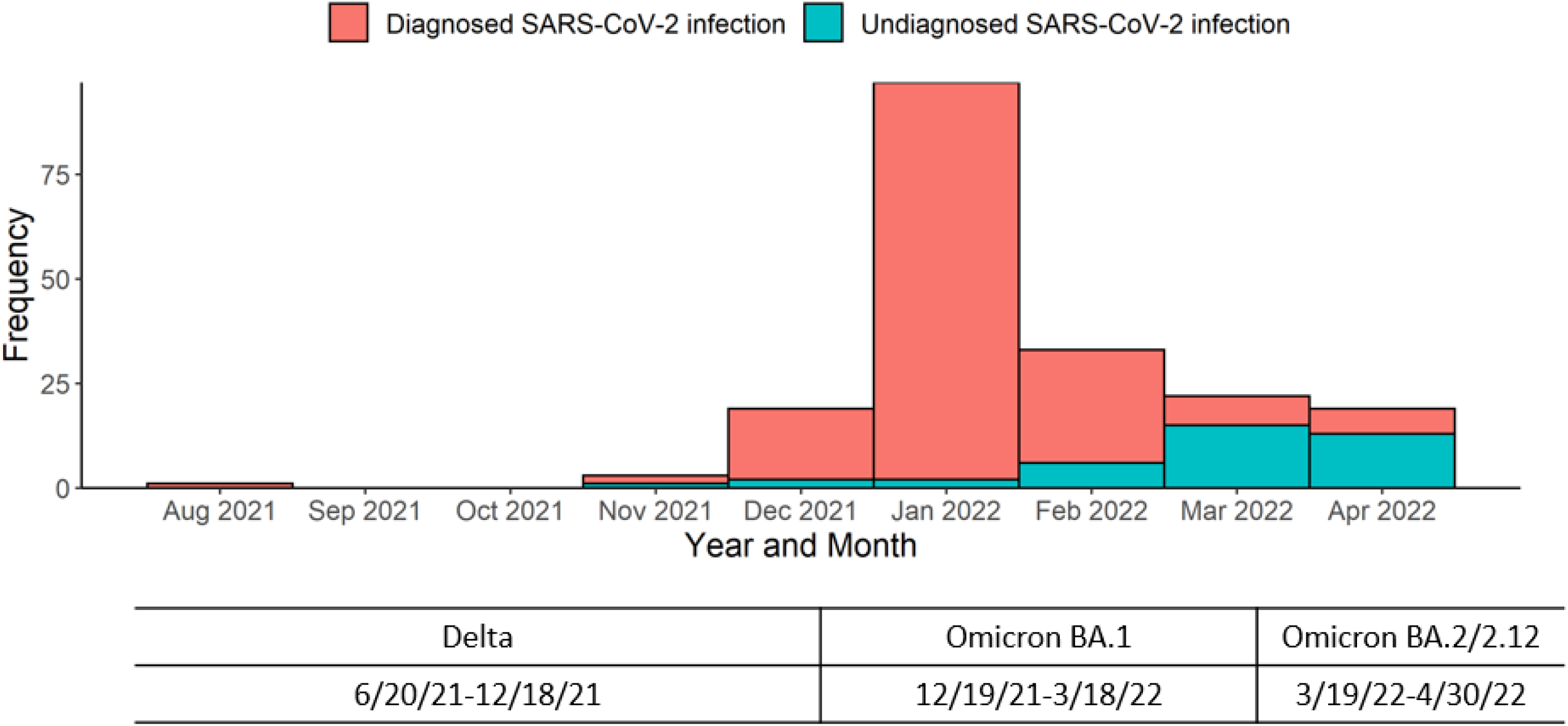
Timing of titer-identified SARS-CoV-2 infections, third dose cohort.

**Table 3.**
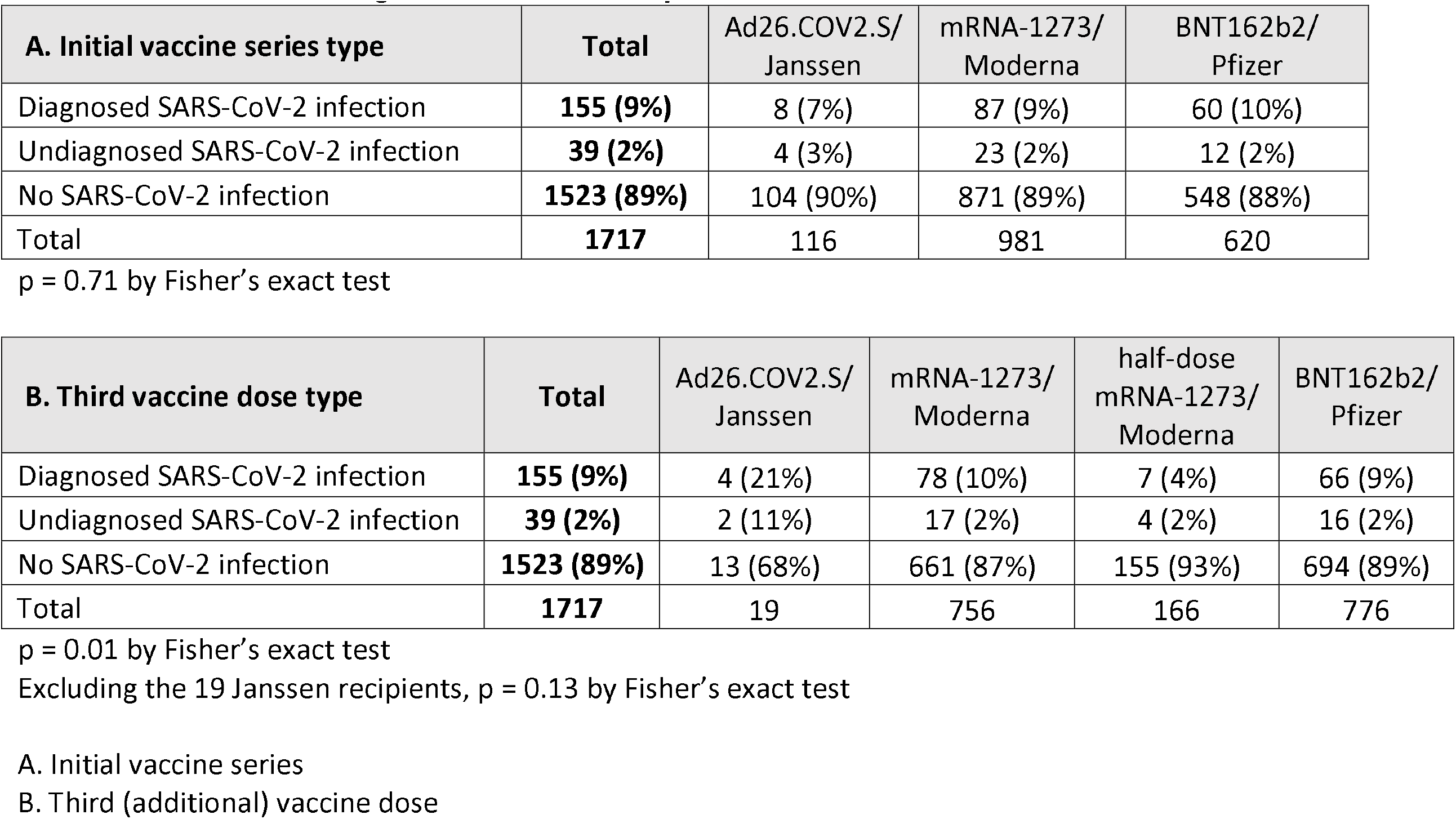
Association of vaccine type with diagnosed and undiagnosed SARS-CoV-2 infections within the third dose cohort among those without history of COVID-19.

| **A. Initial vaccine series type** | **Total** | Ad26.COVS.S/<br>Janssen | mRNA-1273/<br>Moderna | BNT162b2/<br>Pfizer |
| --- | --- | --- | --- | --- |
| Diagnosed SARS-CoV-2 infection | **155 (9%)** | 8 (7%) | 87 (9%) | 60 (10%) |
| Undiagnosed SARS-CoV-2 infection | **39 (2%)** | 4 (3%) | 23 (2%) | 12 (2%) |
| No SARS-CoV-2 infection | **1523 (89%)** | 104 (90%) | 871 (89%) | 548 (88%) |
| Total | **1717** | 116 | 981 | 620 |

p = 0.71 by Fisher's exact test
| **B. Third vaccine dose type** | **Total** | Ad26.COVS.S/<br>Janssen | mRNA-1273/<br>Moderna | half-dose<br>mRNA-1273/<br>Moderna | BNT162b2/<br>Pfizer |
| --- | --- | --- | --- | --- | --- |
| Diagnosed SARS-CoV-2 infection | **155 (9%)** | 4 (21%) | 78 (10%) | 7 (4%) | 66 (9%) |
| Undiagnosed SARS-CoV-2 infection | **39 (2%)** | 2 (11%) | 17 (2%) | 4 (2%) | 16 (2%) |
| No SARS-CoV-2 infection | **1523 (89%)** | 13 (68%) | 661 (87%) | 155 (93%) | 694 (89%) |
| Total | **1717** | 19 | 756 | 166 | 776 |

p = 0.01 by Fisher's exact test
Excluding the 19 Janssen recipients, p = 0.13 by Fisher's exact test
A. Initial vaccine series
B. Third (additional) vaccine dose

Antibody titers were tracked over time following the third vaccine dose. Patients who received Janssen vaccine as the third dose were excluded due to small sample size (N = 19). Antibody titers waned over time among those without a prior history of COVID-19 (**Figure 3B)**. At least 75% of patients had antibody titers at the assay’s upper limit of ≥ 1781 BAU/mL during the first month after receiving the third vaccine dose, regardless of vaccine type. By month 6, median [IQR] titer levels had waned to 1266 [515, ≥ 1781] among Moderna vaccine recipients and 1291 [518, ≥ 1781] among Pfizer vaccine recipients. This trend did not notably differ across types. Of note, the waning is only observable because of the increased assay upper limit, and 76% of patients still have an antibody titer of at least 500 BAU/mL at six months after the third dose.

## Discussion

In this study of patients receiving maintenance dialysis, the first major finding is that a substantial proportion of patients had undiagnosed SARS-CoV-2 infection, comprising 24% and 20% of all SARS-CoV-2 infections, respectively, in the initial vaccine series cohort and the third (additional) dose subcohort, among patients without history of COVID-19 prior to vaccination. The second major finding of this study is that anti-spike IgG antibody titers were higher and waned more slowly after a third vaccine dose than after an initial vaccine series; following an initial vaccine series, 77% of IgG titer levels had waned to ≤ 500 BAU/mL by six months; in contrast, among those who received a third dose, only 24% of titer levels were ≤ 500 BAU/mL at six months.

It is notable that, even among a population of patients with frequent healthcare contact and high utilization, one out of every four to five SARS-CoV-2 infections was undiagnosed. Such “missed” infections likely had only minimal or mild symptoms, and the ability of vaccines to prevent severe COVID-19 (defined by hospitalization for COVID-19 or death) is critical to highlight. At the same time, failure to recognize all SARS-CoV-2 infections remains concerning in maintenance dialysis patients. As medically vulnerable patients who frequently receive care in mandatory congregate settings with high utilization of shared transportation and a high prevalence of long-term care facility residence, the dialysis population remains susceptible to this highly transmissible infection. Other studies on unrecognized SARS-CoV-2 infections either used data from before widespread vaccination of maintenance dialysis patients^17–19^ or were conducted in other populations with different risk factors.^20–22^ These findings highlight the need for ongoing vigilance against COVID-19 as well as maintenance of sensible precautions, such as mask utilization, in patient care areas within dialysis facilities.^23^

Antibody levels were higher among recipients of third vaccine doses beyond the initial series, reinforcing the finding that additional vaccine doses bolster the immune response among maintenance dialysis patients.^7,8,15,24^ However, longer-term studies following the immune response after third doses have so far been limited by sample size and infrequent assessment.^11,12^ This study used six months of post-third dose data to show that third doses also improved durability of the immune response. Specifically, among those without history of COVID-19, median [IQR] titer was 150 [42, 471] BAU/mL at six months in the initial series cohort, similar to our prior findings,^4,13^ but 1288 [512, ≥ 1781] BAU/mL at six months in the third dose cohort. Thus, a three-dose mRNA vaccine series appears superior to a two-dose mRNA vaccine series as a primary vaccine series for patients receiving maintenance dialysis. While the CDC has recommended a three-dose initial series for immunocompromised patients, dialysis patients are not clearly categorized as immunocompromised.

Nevertheless, anti-spike IgG antibody titers waned over time, even after an additional vaccine dose, suggesting a need for subsequent administration of boosters. Serial antibody monitoring among dialysis patients could be explored, both to identify high risk patients who may benefit from additional antiviral strategies for COVID-19 as well as to optimally time future vaccine doses.^24^

This study has several limitations. The assays had upper limits that varied during the study. However, we used multiple methods, including sensitivity analyses, to address this limitation. Second, increases in antibody titer may have occurred due to delayed response to a vaccine dose or lab error. We attempted to account for these by excluding increases around the time of vaccine administration and by using a three-month rolling average, a strategy that does increase the risk of underestimating the number of undiagnosed SARS-CoV-2 infections. We are also unable to identify those with an increase in titer occurring above the assay’s limit of detection, also potentially underestimating undiagnosed infections.

The undiagnosed infections identified in this study occurred during the Delta and Omicron dominant periods, but subsequent patterns of testing and newer Omicron subvariants may impact the future applicability of these results. Lastly, the differences by vaccine type are primarily hypothesis-generating since there may have been confounding by geographic distribution.

In conclusion, among patients receiving maintenance dialysis, there was a high rate of undiagnosed SARS-CoV-2 infections, which supports a role for ongoing common sense precautions, such as mask utilization in dialysis facilities. Higher titer levels and greater durability of titers after a third dose indicate that, among dialysis patients, a three-dose mRNA series is likely the optimal primary series. Moreover, we observed that even after an additional vaccine dose, titers still wane over time, and serial re-dosing of vaccine is likely needed in this vulnerable population.

## Data Availability

These data are not available for sharing.

## Notes

**Support:** This report was supported by Dialysis Clinic, Inc. CMH has received support from ASN KidneyCure’s Ben J. Lipps Research fellowship and from NIH/NCATS grant KL2TR002545. CMH’s funders had no role in study design, data collection, reporting, or the decision to submit.

**Financial Disclosure:** Dr. Manley, Mr. Ladik, Dr. Frament, Dr. Johnson and Dr. Lacson Jr are all employees of DCI, where Dr. Johnson is Vice Chair of the Board. Dr. Weiner, Dr. Miskulin, Dr. Argyropoulos, Dr. Abreo, Dr. Chin, Dr. Gladish, and Dr. Salman receive salary support to their institution from DCI.

### Competing Interest Statement

The authors have declared no competing interest.

### Funding Statement

This report was supported by Dialysis Clinic, Inc. CMH has received support from ASN KidneyCure's Ben J. Lipps Research fellowship and from NIH/NCATS grant KL2TR002545. CMH's funders had no role in study design, data collection, reporting, or the decision to submit.
Dr. Manley, Mr. Ladik, Dr. Frament, Dr. Johnson and Dr. Lacson Jr are all employees of DCI, where Dr. Johnson is Vice Chair of the Board. Dr. Weiner, Dr. Miskulin, Dr. Argyropoulos, Dr. Abreo, Dr. Chin, Dr. Gladish, and Dr. Salman receive salary support to their institution from DCI.

### Author Declarations

This study was reviewed and approved by the WCG IRB Work Order 1-1456342-1.

