## Supplemental Methods for "Serial SARS-CoV-2 antibody titers in vaccinated dialysis patients: prevalence of unrecognized infection and duration of seroresponse"

***Missing and aberrant values***

The SARS-CoV-2 vaccine monitoring protocol automatically deactivates after two consecutive months with SAb-IgG titer less than 1 Index (equivalent to 45 BAU/mL). However, to avoid thereby selecting for patients with better seroresponse, subsequent monthly SAb-IgG levels among patients with consecutive months with SAb-IgG titer below 1 Index were adjudicated as 0 Index (0 BAU/mL) if the patient had not died, received transplantation, developed COVID-19, received a third (additional) vaccine dose, or transferred out of the DCI system. This adjudication is consistent with the assumption behind the protocol's deactivation, specifically that patients who do not have seroimmunity for two consecutive months are unlikely to develop or re-develop it thereafter without exposure to SARS-CoV-2 or undocumented third vaccine dose. There were 905 test results adjudicated this way.

***Defining a threshold to represent Ab-identified SARS-CoV-2 infection***

Excepting assay variability, we do not expect increases in titer except 1) in response to a dose of SARS-CoV-2 vaccine, or 2) in response to SARS-CoV-2 infection. The increases in the 3-month rolling average titer associated with test-diagnosed SARS-CoV-2 infection formed the distribution shown below. Based on the observed distribution and being more than five times the assay's coefficient of variation (17 BAU/mL), we felt that 100 BAU/mL was an acceptable threshold to use to identify SARS-CoV-2 infections, and sensitivity analyses were conducted using 200 BAU/mL as a threshold.

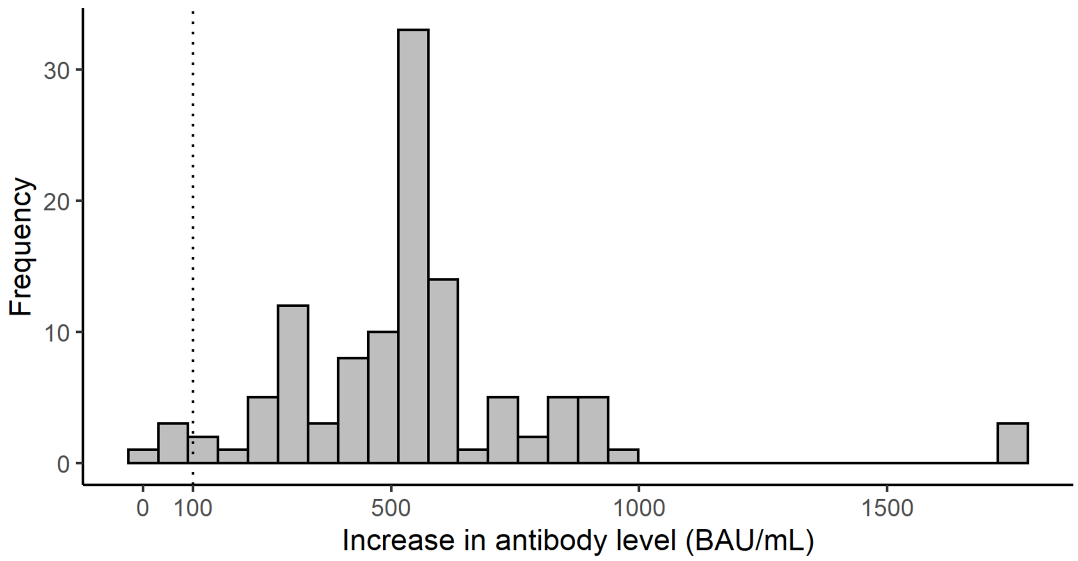


**Figure. Distribution of the increase in 3-month rolling antibody level associated with a test-diagnosed SARS-CoV-2 infection**. Excludes increases of <17 BAU/mL, the assay's coefficient of variation.

***Adjudication of values at the upper limit of the COV2G assay, in the third dose cohort***

As noted, the COV2G assay was used until September 30, 2021, and its upper limit was ≥837 BAU/mL; the manufacturer-updated sCOVG assay was used beginning October 1, 2021 and has an upper limit of ≥1781 BAU/mL. In analyses of the third dose cohort, only 59 values after the third vaccine dose were assessed with the COV2G assay and found to be at its upper limit. Moreover, of values greater than or equal to 837 BAU/mL which were quantified with the sCOVG assay, the median [IQR] value was ≥1781 [≥1781, ≥1781] BAU/mL. Therefore, values which were at the upper limit of the COV2G assay were adjudicated with the value of ≥1781 BAU/mL.
