## Supplemental Tables for "Serial SARS-CoV-2 antibody titers in vaccinated dialysis patients: prevalence of unrecognized infection and duration of seroresponse"

| | Initial series cohort | | | | Third dose cohort | | | |
|---|---|---|---|---|---|---|---|---|
| **Threshold for defining undiagnosed infection by an increase in titer** | **100 BAU/mL** | | **200 BAU/mL** | | **100 BAU/mL** | | **200 BAU/mL** | |
| | n (%) | % of infections | n (%) | % of infections | n (%) | % of infections | n (%) | % of infections |
| Diagnosed SARS-CoV-2 infection | 371 (14%) | 76% | 379 (14%) | 82% | 155 (9%) | 80% | 155 (9%) | 87% |
| Undiagnosed SARS-CoV-2 infection | 115 (4%) | 24% | 85 (3%) | 18% | 39 (2%) | 20% | 23 (1%) | 13% |
| No SARS-CoV-2 infection | 2174 (82%) | --- | 2196 (83%) | --- | 1523 (89%) | --- | 1539 (90%) | --- |
| Total | 2660 | | 2660 | | 1717 | | 1717 | |

**Supplemental Table 1**. Identification of undiagnosed SARS-CoV-2 infections by increase in anti-spike IgG titer. An increase of 100 BAU/mL in the 3-month rolling average titer was used as the primary analysis, and an increase of 200 BAU/mL was used in a sensitivity analysis.

| No prior COVID-19 | | | | | | Prior COVID-19 | | | | | |
|---|---|---|---|---|---|---|---|---|---|---|---|
| | | Initial series | | | | | | Initial series | | | |
| | | J | MM | PP | Total | | | J | MM | PP | Total |
| Third dose | J | 18 | 0 | 1 | 19 | Third dose | J | 5 | 0 | 0 | 5 |
| | M | 25 | 640 | 91 | 756 | | M | 1 | 124 | 15 | 140 |
| | M/2 | 49 | 111 | 6 | 166 | | M/2 | 6 | 11 | 0 | 17 |
| | P | 24 | 230 | 522 | 776 | | P | 6 | 29 | 91 | 126 |
| | Total | 116 | 981 | 620 | 1717 | | Total | 18 | 164 | 106 | 288 |

**Supplemental Table 2**. Among those who received a third (additional) vaccine dose, the breakdown of vaccine type received, by initial vaccine series and by third dose. Here, each capital letter (J, M, or P) represents a single dose of Janssen, Moderna, or Pfizer vaccine, respectively. M/2 represents half-dose Moderna.
