## Supplemental Figures for "Serial SARS-CoV-2 antibody titers in vaccinated dialysis patients: prevalence of unrecognized infection and duration of seroresponse"

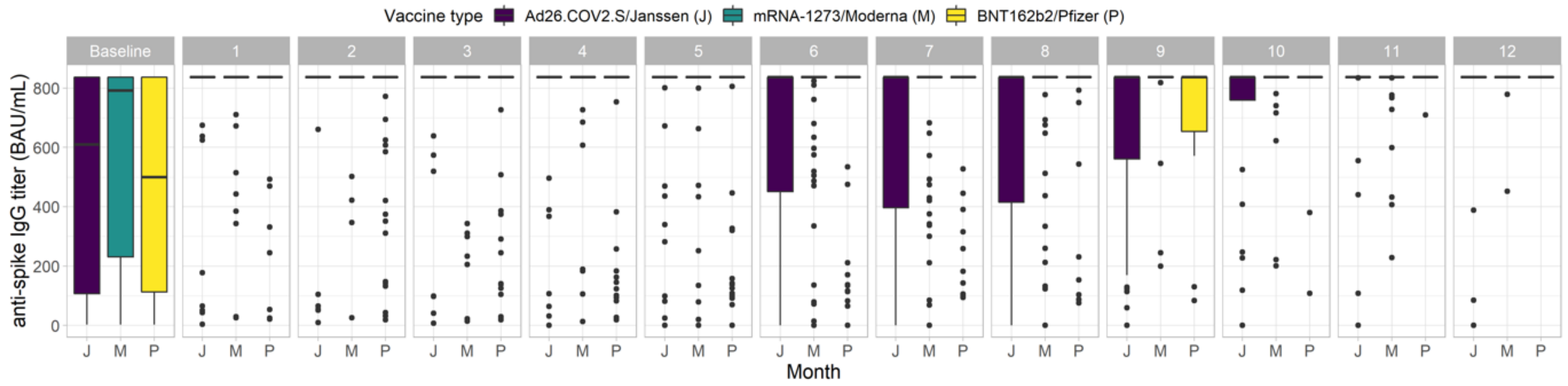


| N = | | | | | | | | | | | | | |
|---|---|---|---|---|---|---|---|---|---|---|---|---|---|
| JANSSEN | 21 | 56 | 58 | 56 | 55 | 57 | 56 | 58 | 49 | 40 | 32 | 26 | 23 |
| MODERNA | 133 | 160 | 177 | 177 | 168 | 144 | 105 | 103 | 69 | 48 | 42 | 34 | 22 |
| PFIZER | 72 | 115 | 108 | 105 | 108 | 90 | 79 | 54 | 37 | 22 | 17 | 18 | 13 |

**Supplemental Figure 1. Anti-spike IgG titers vs months after date of full immunization, comparing by vaccine type, among patients with prior COVID-19 in the initial series cohort**

Of note, the boxplots shown here are bounded by the upper and lower limits of the assay used. The dots represent the outliers, defined as greater than 1.5*IQR above the third quartile or less than 1.5*IQR below the first quartile.

The tables of N show the number of titers for each month, by vaccine type.

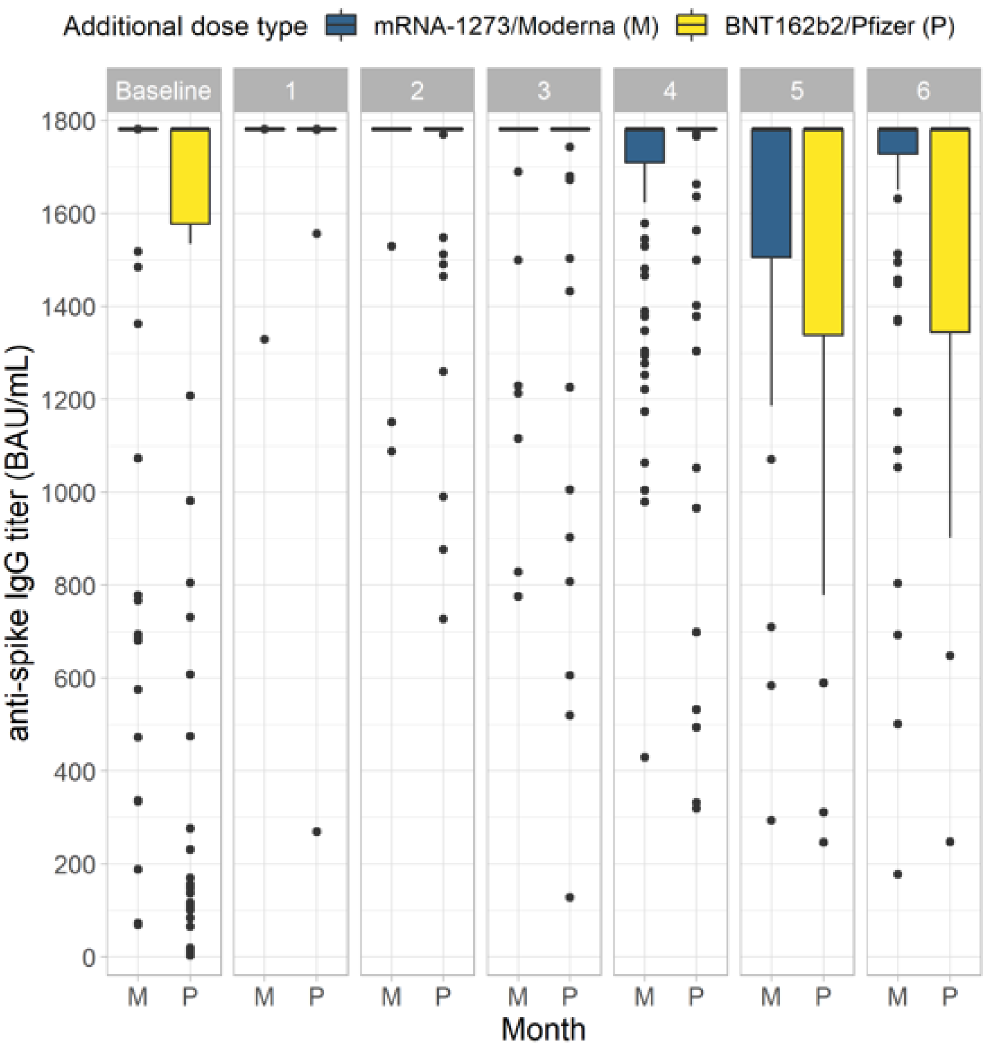


| N = | | | | | | | |
|---|---|---|---|---|---|---|---|
| MODERNA | 115 | 103 | 104 | 105 | 90 | 71 | 68 |
| PFIZER | 95 | 91 | 93 | 80 | 67 | 38 | 47 |

**Supplemental Figure 2. Anti-spike IgG titers vs months after date of full immunization by third vaccine dose, comparing by vaccine type, among patients with prior COVID-19 in the third dose cohort**

Of note, the boxplots shown here are bounded by the upper and lower limits of the assay used. The dots represent the outliers, defined as greater than 1.5*IQR above the third quartile or less than 1.5*IQR below the first quartile.

The tables of N show the number of titers for each month, by vaccine type.
